# Chest Radiography AI Concordance and Lung Cancer Linkage in a Large Health Check-up Cohort

**DOI:** 10.64898/2026.07.27.26359077

**Authors:** Yu Fujita, Susumu Saito, Shigehiro Yagishita, Jun Araya, Ryo Nakagawa

## Abstract

**Purpose:** To evaluate the implementation characteristics of a commercially available chest radiography artificial intelligence (AI) system in a large real-world health check-up cohort using workflow-level, lesion-specific, and exploratory retrospective lung cancer case analyses.

**Methods:** This retrospective single-centre study included 298,991 consecutive health check-up chest radiographs from 114,866 individuals obtained between 2019 and 2023 and interpreted under routine double reading by board-certified radiologists. A commercially available AI system was evaluated using two prespecified thresholds: positivity in any of ten findings for the all-score analysis and positivity for nodule or mass for the nodule-focused analysis, both at a manufacturer-recommended score threshold of 15. Because routine radiologist judgement rather than universal CT or pathologic verification served as the reference framework, the primary analyses were interpreted as radiologist-referenced operational concordance analyses.

**Results:** Radiologist-referenced sensitivity and specificity were 72.0% and 79.6%, respectively, in the all-score analysis and 87.1% and 91.8%, respectively, in the nodule-focused analysis. Negative predictive values were 99.0% and 100.0%, respectively. Among 48 histopathologically confirmed lung cancer cases, retrospective timeline analyses showed earlier AI positivity than routine radiologist positivity in a subset of cases. These findings should be interpreted as exploratory observations and do not establish prospective clinical benefit.

**Conclusion:** In a large health check-up cohort, chest radiography AI demonstrated stable concordance with routine radiologist judgement and high sensitivity for radiologist-reported pulmonary nodules and masses. Exploratory retrospective analyses showed earlier AI positivity in a subset of histopathologically confirmed lung cancer cases, supporting further prospective evaluation of AI-assisted health check-up workflows.

**At a Glance Commentary:** *Scientific Knowledge on the Subject:* Artificial intelligence has shown promise for supporting chest radiograph interpretation, including detection of pulmonary nodules and other thoracic abnormalities. However, most previous studies have used selected datasets, simulated reading environments, or enriched populations. Evidence remains limited regarding how chest radiography AI performs at programme scale in low-prevalence, real-world health check-up settings, particularly with respect to concordance with routine radiologist judgement, discordant review volume, and retrospective linkage to clinically confirmed lung cancer cases.

*What This Study Adds to the Field:* In 298,991 consecutive health check-up examinations, chest radiography AI demonstrated stable concordance with routine radiologist judgement. For radiologist-reported pulmonary nodules and masses, AI showed 87.1% sensitivity and 100.0% negative predictive value relative to the radiologist reference framework. Retrospective analysis of histopathologically confirmed lung cancer cases identified earlier AI positivity in a subset of patients, although prospective clinical benefit remains unproven. These findings provide large-scale, implementation-oriented evidence on the potential role and limitations of AI as a supplementary tool within radiologist-led chest radiography workflows.

## Introduction

Low-dose CT screening reduces lung cancer mortality in selected high-risk populations and established the principle that imaging-based screening can improve outcomes. The National Lung Screening Trial showed a 20.0% reduction in lung cancer mortality with low-dose CT versus chest radiography, and the NELSON trial subsequently confirmed a significant mortality reduction with volume CT screening in a European randomised population (1, 2). These trials, together with more recent guideline updates, provide the foundation for contemporary lung cancer screening strategies, but broad implementation remains constrained by imaging capacity, programme infrastructure, eligibility determination, and downstream testing burden (1–4). However, chest radiography itself has not demonstrated a significant lung cancer mortality benefit in large randomised trials such as PLCO (5). Therefore, the present study should not be interpreted as an evaluation of chest radiography as a substitute for LDCT-based lung cancer screening, but rather as an implementation-oriented evaluation of AI-assisted decision support in routine health check-up chest radiography.

AI for chest radiography has advanced rapidly in recent years. Previous studies have shown that deep learning systems can support abnormality triage, improve radiologist performance across multiple thoracic findings, and identify clinically relevant missed findings within routine workflows (6–11). However, most investigations have focused on selected tasks, enriched datasets, or simulated reading environments. Evidence remains limited regarding the operational consequences of fixed-threshold AI deployment in large real-world health check-up cohorts.

Despite this progress, evidence remains limited for large, unselected health check-up cohorts evaluated within routine workflows. Many previous studies were conducted in enriched or experimental settings, focused on selected findings, or assessed simulated worklists rather than real programme-scale operational consequences. In particular, few studies have combined three implementation-relevant components within a single real-world health check-up cohort: broad abnormality triage, lesion-specific nodule assessment, and retrospective linkage to confirmed lung cancer cases. This gap is especially important in low-prevalence settings, where operational consequences such as positivity rate, discordant review volume, and escalation workload may be as important as conventional performance metrics.

In this study, we evaluated a commercially available chest radiography AI system in a large consecutive health check-up cohort interpreted under routine double reading. The primary objective was to assess workflow-level concordance between AI positivity and routine radiologist judgement using the maximum score across ten findings. Secondary objectives were to assess lesion-specific concordance for pulmonary nodules and masses and to perform an exploratory retrospective linkage analysis among confirmed lung cancer cases. By explicitly separating workflow concordance, lesion-focused agreement, and outcome-linked timeline assessment, this study was designed to evaluate the implementation-relevant role of AI in routine chest radiography without overextending the results to claims of autonomous lung cancer screening efficacy.

## Materials and Methods

### Study design and reporting framework

We conducted a retrospective single-centre observational study at Omiya City Clinic, Japan, using consecutive health check-up chest radiography examinations performed between Jan 1, 2019, and Dec 31, 2023. The study was designed as an implementation-oriented evaluation of AI performance against routine radiologist judgement embedded in a real-world workflow. Reporting was informed by STROBE, STARD 2015, and STARD-AI, with prespecification of the index test definition, reference framework, and primary operational metrics (12–14). Because the primary reference framework was routine radiologist judgement rather than a universal CT- or pathology-based outcome standard, the main cohort analyses were designed to assess implementation-relevant concordance with real-world workflow rather than absolute disease detection accuracy.

### Routine imaging and reading workflow

Chest radiographs were acquired as part of standard health check-up practice. Routine interpretation followed the established local workflow, including double reading by board-certified radiologists. Routine judgement was recorded using the standard Japanese health check-up category system for imaging findings, based on the standardized reporting framework recommended by the Japanese Society of Ningen Dock and Preventive Medical Care (15), in which category A denotes no abnormality, category B minor abnormality, category C repeat examination or interval follow-up, and category D detailed diagnostic evaluation or treatment. Routine abnormality was defined as any category other than A or B. Higher-acuity subcategories were grouped as abnormal. This pragmatic reference framework was used to evaluate interaction with real-world workflow rather than disease-specific sensitivity (13, 14).

### Data linkage

Imaging data, AI outputs, and routine judgements were linked using anonymised identifiers and examination date. For the lung cancer timeline analysis, confirmed lung cancer cases diagnosed between 2019 and 2023 were identified from clinic records and available diagnostic information. All lung cancer cases included in the timeline analysis were histopathologically confirmed by biopsy or surgical resection. Historical screening timelines were reconstructed using screening dates, AI outputs, routine judgements, and chest CT where available. This analysis was defined as an exploratory retrospective timeline linkage analysis, not as a prospective screening outcome analysis or proof of earlier clinical cancer detection. Extracted variables included age, sex, screening date, AI and radiologist positivity, first positive dates, diagnosis date, detection category, and lead time. Diagnostic subtype and TNM stage (9th edition) were recorded where available.

## Results

### Cohort selection and patient characteristics

The source dataset contained 299,745 screening chest radiography examinations. Of these, 299,712 were successfully linked to AI outputs, and 298,991 examinations from 114,866 patients remained after exclusion of 721 examinations with missing values in one or more AI score fields (**Figure 1**). By year, the final included examinations numbered 60,363 in 2019, 52,757 in 2020, 60,769 in 2021, 62,092 in 2022, and 63,010 in 2023 (**sTable 1**). Baseline characteristics of the analytical cohort are summarised in **sTable 1**.

**Figure 1.**
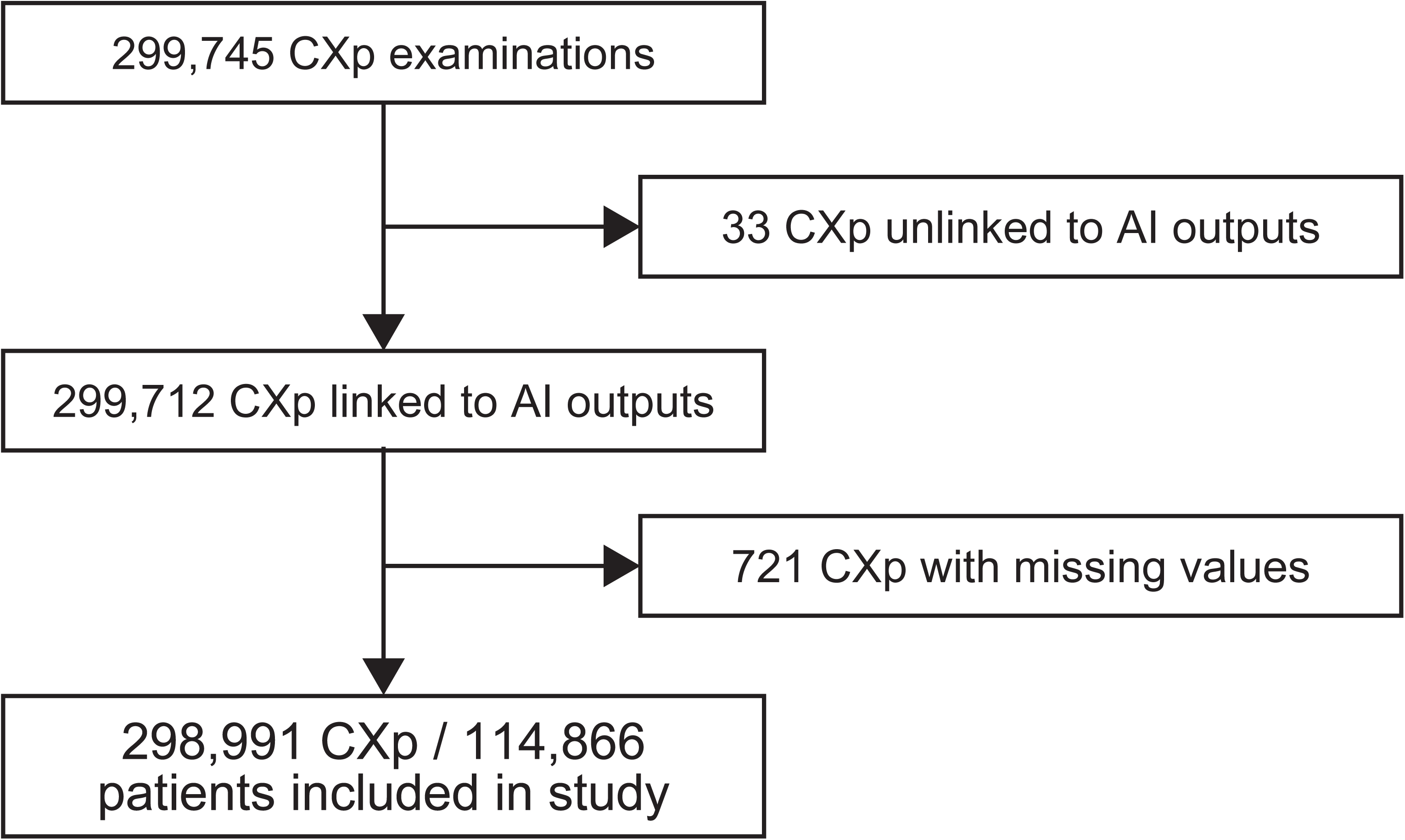
Study selection flow. Flow diagram showing derivation of the final analytical cohort from the source screening chest radiography dataset, including linkage to AI outputs and exclusion of examinations with missing AI scores.

### Primary analysis: workflow-level concordance using the all score

In the all-score analysis, AI was positive in 65,303 of 298,991 examinations, whereas routine abnormality was recorded in 8,293 examinations. Relative to routine radiologist judgement, the overall 2 × 2 table comprised 5,975 AI-positive/routine-abnormal examinations, 59,328 AI-positive/routine-normal examinations, 2,318 AI-negative/routine-abnormal examinations, and 231,370 AI-negative/routine-normal examinations (**Figure 2A**, **Table 1**, **sTable 2**). Radiologist-referenced sensitivity was 72.0%, specificity 79.6%, PPV 9.1%, and NPV 99.0% (**Figure 2A**, **Table 1**).

**Figure 2.**
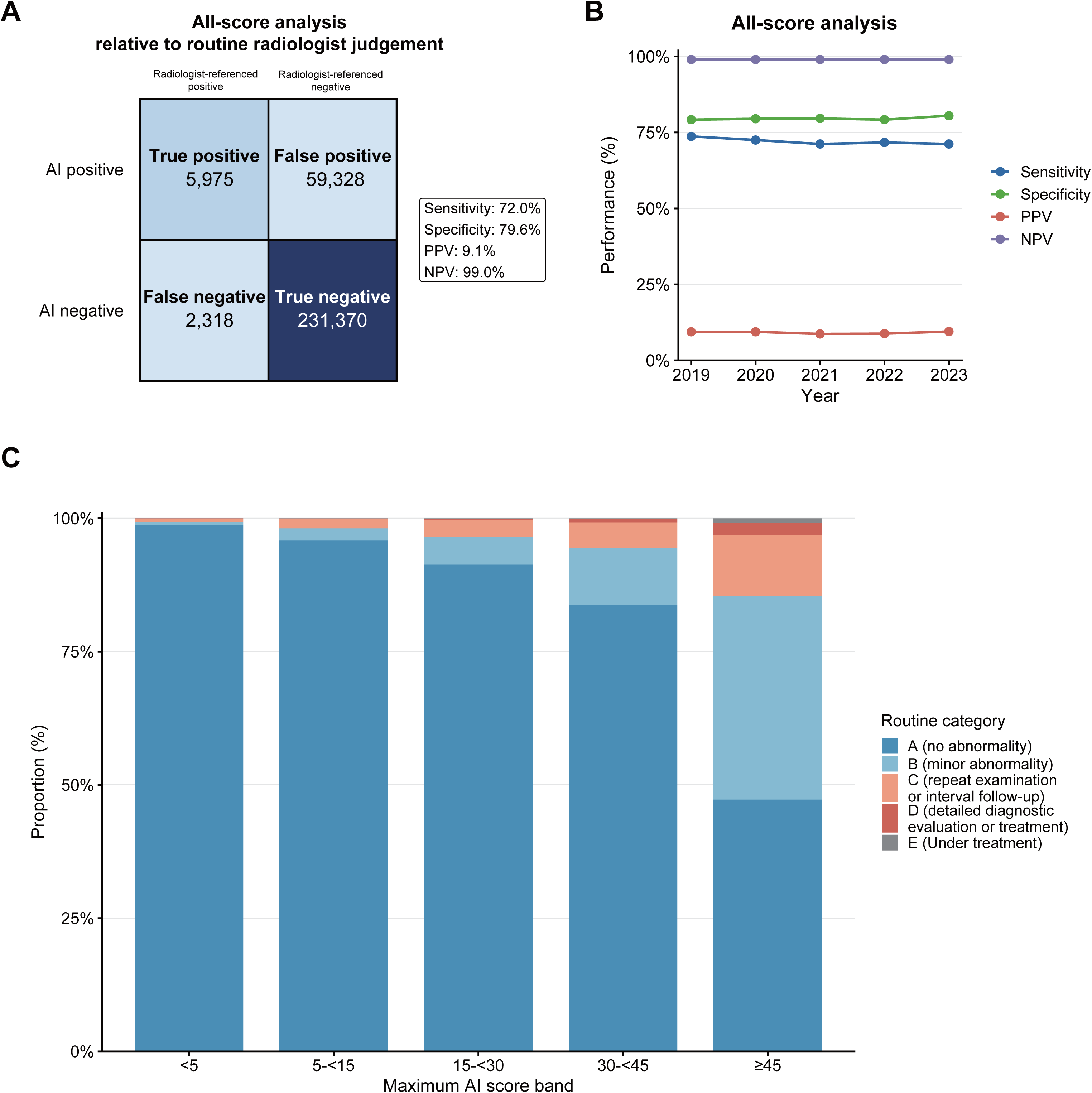
Primary all-score analysis: workflow-level concordance and enrichment across maximum AI score bands. (A) Confusion matrix for the primary all-score analysis using routine radiologist judgement as the reference framework. (B) Year-by-year radiologist-referenced concordance metrics for the all-score analysis. (C) Stacked distribution of routine screening categories according to maximum AI score band. In the all-score analysis, AI positivity was defined as a score of 15 or higher in any of ten thoracic abnormality categories, and the reference framework was routine radiologist judgement of abnormality defined as any screening category other than A or B. The confusion matrix shows concordant-positive, discordant AI-positive, discordant AI-negative, and concordant-negative examinations relative to routine judgement. The line plot shows annual sensitivity, specificity, PPV, and NPV from 2019 to 2023. For panel C, examinations were stratified by the highest score across the ten thoracic abnormality categories (<5, 5 to <15, 15 to <30, 30 to <45, and ≥45), and the proportions of routine screening categories A–E are shown within each band. PPV; positive predictive value, NPV; negative predictive value.

**Table 1.** Diagnostic performance relative to routine radiologist judgement.

| Analysis | TP | FP | FN | TN | Sensitivity (%; 95% CI) | Specificity (%; 95% CI) | PPV (%; 95% CI) | NPV (%; 95% CI) |
| --- | --- | --- | --- | --- | --- | --- | --- | --- |
| All-score | 5,975 | 59,328 | 2,318 | 231,370 | 72.0 (70.4–73.5) | 79.6 (79.3–79.8) | 9.1 (8.8–9.5) | 99.0 (98.9–99.1) |
| Nodule-focused | 398 | 24,402 | 59 | 274,132 | 87.1 (84.1–90.5) | 91.8 (91.7–92.0) | 1.6 (1.4–1.8) | 100.0 (100.0–100.0) |

In the all-score analysis, AI positivity was defined as a score of 15 or higher in any of ten AI output categories, and the reference framework was routine radiologist judgement of abnormality defined as categories other than A or B. In the nodule-focused analysis, AI positivity was defined as a nodule or mass score of 15 or higher, and the reference framework was radiologist-reported pulmonary nodule or mass findings. TP, FP, FN, and TN were defined relative to the predefined radiologist reference framework and do not necessarily represent pathologically confirmed disease status. TP; true-positive, FP; false-positive, FN; false-negative, TN; true-negative, PPV; positive predictive value, NPV; negative predictive value, CI; confidence intervals.

Year-by-year performance in the all-score analysis was stable across the study period (**Figure 2B**, **sTable 1**, **sTable 2**, **sTable 3**). Sensitivity ranged from 71.2% to 73.7%, specificity from 79.2% to 80.5%, PPV from 8.7% to 9.5%, and NPV remained 99.0% in every year.

Score-band analysis showed a progressive shift in routine screening categories as the maximum AI score increased (**Figure 2C**). In examinations with maximum AI scores below 5, category A accounted for 98.77% of cases. By contrast, in examinations with scores of 45 or higher, category A decreased to 47.24%, whereas category B increased to 38.15% and higher-acuity categories became more frequent. These findings indicate that the all-score was associated with routine radiologist judgement categories, but they should not be interpreted as formal calibration of disease probability.

Among AI-positive but routine-normal examinations in the all-score analysis, 59,328 examinations were classified as discordant AI-positive examinations relative to routine judgement. The most common routine label within this group was no abnormality, accounting for 45,111 examinations (76.0%), followed by minor abnormality in 14,217 examinations (24.0%; **sTable 4**, **sFigure 1**). Among radiologist-reported findings, no abnormality was the most frequent category, followed by old inflammatory changes, atelectasis, and pleural thickening (**sTable 4**, **sFigure 1**). These findings objectively describe the potential additional review volume that would arise if AI positivity were used as a screening escalation trigger.

### Secondary analysis: nodule-focused concordance

In the nodule-focused analysis, 457 of 298,991 examinations were radiologist-positive for pulmonary nodules or masses, whereas 298,534 were radiologist-negative. Relative to the radiologist-defined nodule or mass reference framework, the AI nodule or mass score yielded 398 concordant-positive, 24,402 discordant AI-positive, 59 discordant AI-negative, and 274,132 concordant-negative examinations (**Figure 3A**, **Table 1**, **sTable5**). Radiologist-referenced sensitivity was 87.1%, specificity 91.8%, PPV 1.6%, and NPV 100.0% (**Figure 3A**, **Table 1**).

**Figure 3.**
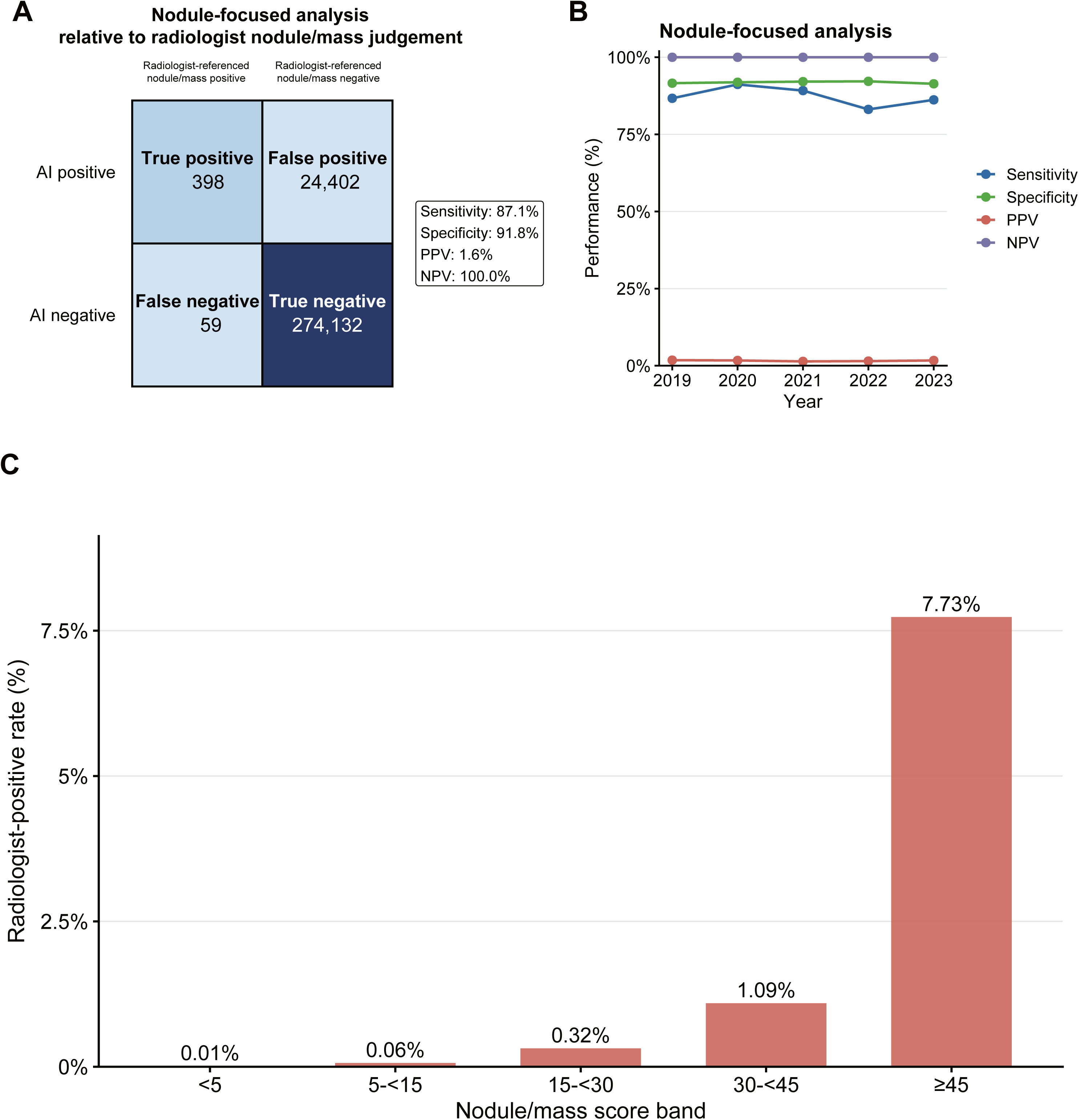
Secondary nodule-focused analysis: lesion-specific concordance and enrichment across nodule or mass AI score bands. (A) Confusion matrix for the secondary nodule-focused analysis using radiologist-reported pulmonary nodule or mass findings as the reference framework. (B) Year-by-year radiologist-referenced concordance metrics for the nodule-focused analysis. (C) Radiologist-positive pulmonary nodule or mass rate according to nodule or mass AI score band. In the nodule-focused analysis, AI positivity was defined as a nodule or mass score of 15 or higher, and the reference framework was radiologist-reported pulmonary nodule or mass findings. The confusion matrix shows concordant-positive, discordant AI-positive, discordant AI-negative, and concordant-negative examinations relative to radiologist judgement. The line plot shows annual sensitivity, specificity, PPV, and NPV from 2019 to 2023. For panel C, examinations were stratified by the nodule or mass score (<5, 5 to <15, 15 to <30, 30 to <45, and ≥45), and the proportions of radiologist-positive and radiologist-negative examinations are shown within each band. PPV; positive predictive value, NPV; negative predictive value.

Year-by-year performance in the nodule-focused analysis was also stable (**Figure 3B**, **sTable 5**). Sensitivity ranged from 83.1% to 91.2%, specificity from 91.4% to 92.2%, PPV from 1.4% to 1.8%, and NPV remained 100.0% throughout the study period.

The prevalence of radiologist-positive pulmonary nodules or masses was low in the health check-up cohort. Score-band analysis provided additional operational context (**Figure 3C**). The radiologist-positive rate rose steadily across nodule or mass score bands, reaching 7.73% in examinations with scores of 45 or higher. Thus, higher score bands identified enriched subgroups relative to routine radiologist judgement; however, clinical escalation based on these bands would require threshold sensitivity analysis, assessment of workload trade-offs, and prospective validation.

Among AI-positive but radiologist-negative examinations in the nodule-focused analysis, discordant AI-positive findings were dominated by routine categories A and B, which together accounted for 89.5% of such examinations (**sTable 6**, **sFigure 2**). In the radiologist-reported finding breakdown, no abnormality and old inflammatory changes were the most frequent categories, followed by cyst, post-surgical change, and non-tuberculous mycobacterial disease (**sTable 6**, **sFigure 2**). These findings describe benign mimics and potential additional review volume, but do not prove that all AI-positive/radiologist-negative examinations were biologically false positive.

### Threshold sensitivity analysis

Radiologist-referenced ROC analysis demonstrated good discrimination in both analyses (**sFigure 3**, **sTable 7**). The area under the ROC curve (AUC) was 0.828 (95% CI, 0.823–0.833) for the all-score analysis and 0.950 (95% CI, 0.938–0.962) for the nodule-focused analysis. The prespecified operational threshold of 15 was located close to the ROC-derived Youden-optimal threshold in both analyses. In the all-score analysis, the Youden-optimal threshold was 16.9, yielding a sensitivity of 71.2% and specificity of 80.8%, compared with 72.0% and 79.6%, respectively, at the prespecified threshold of 15. Similarly, in the nodule-focused analysis, the Youden-optimal threshold was 18.1, yielding a sensitivity of 85.3% and specificity of 94.0%, compared with 87.1% and 91.8%, respectively, at the prespecified threshold. These findings indicate that the manufacturer-recommended threshold of 15 operates near the ROC-derived optimum and that modest threshold increases would provide only limited gains in specificity at the expense of sensitivity. Accordingly, the selected threshold appears operationally reasonable for implementation-oriented evaluation within this health check-up cohort.

### Exploratory tertiary analysis: retrospective lung cancer timeline linkage

The linked outcome cohort included 48 confirmed lung cancer cases diagnosed between 2019 and 2023. When classified according to the relationship between AI positivity and radiologist positivity at relevant historical screening timepoints, 20 cases showed simultaneous AI and radiologist positivity, 11 showed earlier AI positivity, 10 showed AI-only positivity before diagnosis, two showed radiologist-only positivity, and five were not positive by either method at the relevant screening timepoint, although these five cases were subsequently identified by optional same-day chest CT (**Figure 4A**, **sTable 8**).

**Figure 4.**
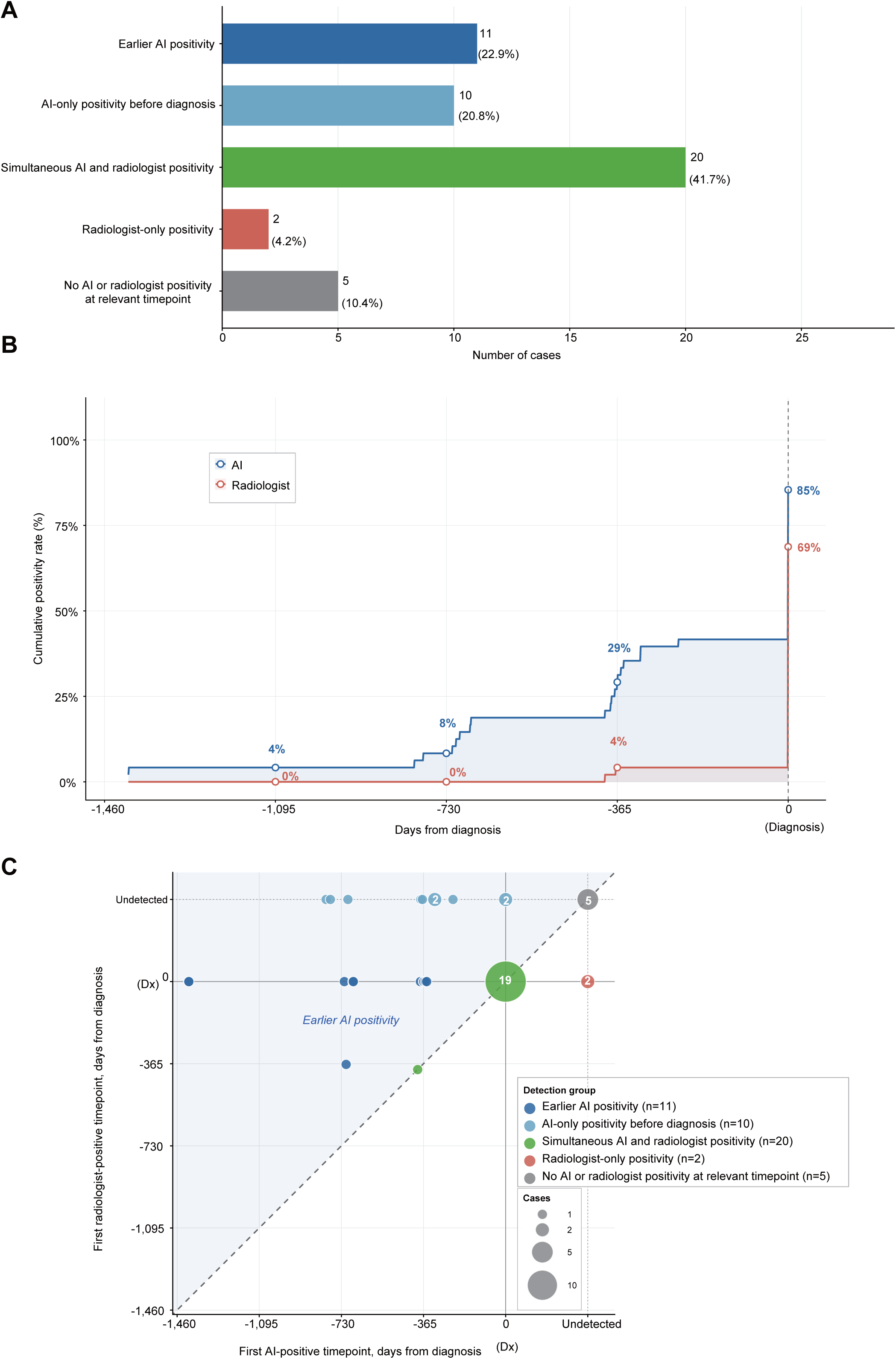
Exploratory retrospective AI positivity timelines among confirmed lung cancer cases. (A) Retrospective positivity patterns among confirmed lung cancer cases. Cases were classified as showing earlier AI positivity, AI-only positivity before diagnosis, simultaneous AI and radiologist positivity, radiologist-only positivity, or no positivity by either method at the relevant screening timepoint. (B) Cumulative retrospective positivity curves for AI and routine radiologist judgement before diagnosis. The x-axis represents days from diagnosis, with negative values indicating time before diagnosis. Curves show the cumulative proportion of cases with AI positivity or radiologist positivity over time. (C) Relationship between the timing of first AI positivity and first radiologist positivity relative to diagnosis. Each bubble represents one or more cases according to the first AI-positive and first radiologist-positive timepoints. Cases without positivity by one modality were plotted in the corresponding undetected category. The diagonal line indicates equal timing of positivity for AI and radiologist judgement.

Cumulative positivity curves showed that AI became positive in a greater proportion of confirmed lung cancer cases than routine radiologist interpretation across selected prediagnostic intervals (**Figure 4B**). AI positivity reached 4% by approximately 1,095 days before diagnosis, 8% by around 730 days, 29% by around 365 days, and 85% by the time of diagnosis, whereas the corresponding cumulative radiologist positivity rates were 0%, 0%, 4%, and 69%, respectively. These curves represent retrospective AI score positivity and should not be interpreted as evidence of prospective clinical benefit.

Lead-time analysis was possible in 41 cases with identifiable dates of first AI positivity and confirmed diagnosis. The mean interval from first AI positivity to diagnosis was 266.7 days, the median was 0 days, and the maximum was 1409 days (**sTable 8**, **sTable 9**), indicating that the mean was influenced by a limited number of cases with long prediagnostic intervals. In 19 cases, AI positivity occurred before radiologist positivity according to the predefined temporal classification framework, whereas no case met the predefined criterion for earlier radiologist positivity than AI (**Figure 4C**).

Case-level timelines illustrated heterogeneous patterns (**sTable 9**). In some cases, AI positivity preceded both radiologist positivity and confirmed diagnosis by months to years, suggesting a potential safety-net signal. In other cases, AI remained negative despite a radiologist-flagged lesion, underscoring the continued importance of expert human interpretation. Because this analysis was retrospective, the findings are hypothesis-generating and are subject to lead-time bias, length-time bias, and possible overdiagnosis of indolent lesions.

## Discussion

In this large real-world health check-up cohort, chest radiography AI showed implementation-relevant associations at three complementary levels, including workflow-level concordance with routine abnormality judgement, lesion-specific concordance for pulmonary nodules and masses, and exploratory retrospective positivity patterns among confirmed lung cancer cases. This three-layer framework extends conventional cross-sectional agreement analyses and addresses practical aspects of AI implementation in health check-up programmes, including positivity rates, discordant review volume, escalation pathways, and the timing of abnormal signals. Importantly, because the main reference framework was routine radiologist judgement rather than universal CT or pathological verification, these findings should be interpreted as evidence of operational concordance within a real-world workflow rather than definitive diagnostic accuracy or proof of lung cancer screening efficacy. Our findings are consistent with prior studies showing that chest radiography AI can support abnormality triage, radiologist performance, and workflow efficiency (6–9, 16). However, most previous studies evaluated reader performance, simulated workload reduction, or selected implementation settings rather than the programme-scale operational consequences of fixed-threshold AI deployment in a real-world health check-up cohort.

Our results emphasise that, in low-prevalence health check-up settings, positivity rates and discordant review volume are central implementation outcomes. In the all-score analysis, AI positivity substantially exceeded the routine abnormality rate, indicating that workflows using AI positivity for additional review or CT escalation would need to absorb substantial downstream workload. These AI-positive/routine-normal examinations should be interpreted as discordant findings rather than definitive false positives, because some may represent true abnormalities not recorded in the routine reference framework. Consistent with real-world imaging AI implementation literature (17), our findings suggest that fixed-threshold AI triage may be more suitable as a safety-net signal than as a workload-reducing gatekeeping strategy at the evaluated operating point.

The nodule-focused analysis is clinically relevant to health check-up chest radiography but remains distinct from validated lung cancer screening performance. Consistent with a randomized trial showing improved detection of actionable lung nodules with AI assistance (10), our nodule or mass analysis showed high radiologist-referenced sensitivity and very high negative predictive value despite the low prevalence of radiologist-positive examinations. Higher score bands also identified enriched subgroups relative to routine radiologist judgement. These findings support targeted second review or prior-image comparison for selected high-score cases, but score-based escalation requires prospective validation and should not be interpreted as calibrated malignancy risk. ROC-based threshold sensitivity analyses further showed that the manufacturer-recommended threshold of 15 was located close to the Youden-optimal operating point in both analyses, supporting its use as a pragmatic prespecified threshold for implementation-oriented evaluation.

The lung cancer timeline analysis adds a clinically relevant but exploratory dimension that is often missing from studies evaluating chest radiography AI. Cross-sectional concordance analyses cannot determine whether abnormal AI signals occur sufficiently early to influence follow-up decisions. In our cohort, some confirmed lung cancer cases demonstrated AI positivity before routine radiologist positivity, whereas others were flagged only by AI before diagnosis. However, these retrospective observations should not be interpreted as evidence of earlier clinically beneficial detection or improved patient outcomes. The median lead time was 0 days, and the mean lead time was influenced by a limited number of cases with long prediagnostic intervals. Furthermore, lead-time bias, length-time bias, and overdiagnosis cannot be excluded in this retrospective analysis. Nevertheless, these findings suggest that AI may provide an additional safety-net signal in selected cases, which is consistent with previous studies showing that AI-assisted secondary review can identify a small but clinically relevant number of missed findings, particularly pulmonary nodules, within established radiologist-led workflows (11). Prospective studies incorporating predefined management pathways are required to determine whether such signals influence interval cancer rates, stage distribution at diagnosis, or patient outcomes.

Taken together, these findings support structured integration of chest radiography AI into radiologist-led health check-up workflows rather than autonomous triage. Potential implementation strategies include second-look safety-net review, worklist prioritisation, and score-based escalation using predefined thresholds. Regardless of the strategy, safeguards are needed to reduce automation bias and to guide management of highly discordant AI-positive/radiologist-negative or AI-negative/radiologist-positive examinations. Because clinical guidelines for such discordant cases remain limited, prospective deployment should include predefined responsibilities, escalation criteria, and documentation procedures.

This study has limitations. First, the main cohort analysis used routine radiologist judgement rather than a universal CT- or pathology-based reference standard, making it most appropriate for evaluating interaction with real workflow rather than absolute disease detection. Consequently, AI-positive/radiologist-negative examinations may include both non-actionable AI findings and true abnormalities missed or not recorded during routine interpretation. Second, this was a single-centre study, and the generalisability of the findings might be influenced by local imaging protocols, reading practices, and population characteristics. Third, detailed reader-level information regarding the double-reading workflow, reader experience, independence of reads, and adjudication of disagreements was not consistently available. Fourth, the lung cancer linkage analysis was descriptive and involved a modest number of confirmed cases, and it is subject to lead-time bias, length-time bias, and possible overdiagnosis. Finally, we evaluated a single prespecified operating threshold; alternative thresholds or score-band strategies could result in different trade-offs between sensitivity, workload, and downstream testing.

Future work should move beyond concordance metrics alone and prospectively test workflow designs in which operating thresholds, escalation criteria, and downstream imaging pathways are specified in advance. Programme-level endpoints such as positivity rate, discordant review volume, CT utilisation, time to diagnosis, interval cancers, stage distribution at diagnosis, and patient outcomes should be treated as core outcomes for health check-up chest radiography AI evaluation. This perspective is consistent with recent reporting guidance for AI-centred diagnostic studies, including STARD-AI, which emphasises transparent prespecification, clear intended use, and applicability to the real clinical pathway (14). In parallel, broader lung cancer screening policy remains anchored in low-dose CT for eligible high-risk individuals (1–3), whereas chest radiography AI may have value as a scalable adjunct in health check-up systems where chest radiography is already deeply embedded. This potential adjunctive role may be most relevant in settings where CT-based screening access is constrained by geography, affordability, or service capacity (4).

In conclusion, in a large low-prevalence health check-up cohort interpreted under routine double reading, chest radiography AI demonstrated stable workflow-level concordance with routine abnormality judgement and high sensitivity for radiologist-reported nodules and masses. In exploratory retrospective analyses, earlier AI positivity was observed in a subset of confirmed lung cancer cases. These findings support further evaluation of AI as a safety-net and prioritisation tool within radiologist-led chest radiography workflows, but they do not support replacement of radiologist review or claims of proven lung cancer screening benefit at the evaluated operating threshold. More broadly, our results indicate that meaningful evaluation of chest radiography AI should include not only concordance with routine judgement, but also programme-scale operational consequences, management of discordant cases, and linkage to clinically relevant outcomes in prospective studies.

## Author contributors

Y.F. and R.N. conceived the study. S.Y. curated the dataset and coordinated data governance. S.S. and J.A. contributed to clinical interpretation of screening workflow and outcome linkage. S.Y. and R.N. performed statistical analyses. Y.F. drafted the manuscript. All authors interpreted the data, critically revised the manuscript for important intellectual content, and approved the final version.

Y.F. had full access to all the data in the study and had final responsibility for the decision to submit for publication.

## Data Availability

All data produced in the present study are available upon reasonable request to the authors.

## Acknowledgments

No specific funding was received for this study. CXR-AID was provided free of charge by FUJIFILM for use during the study period. FUJIFILM had no role in the study design, data collection, data analysis, data interpretation, writing of the report, or the decision to submit the paper for publication.

## Supplementary appendix

**Supplementary Table 1.** Baseline characteristics of the analytical cohort Characteristic Value. Baseline characteristics of the final analytical cohort after linkage of screening chest radiography examinations and Al outputs and exclusion of examinations with missing Al scores. The table summarises cohort size, patient demographics, smoking status, examination frequency per patient, and annual distribution of included examinations.

| Characteristic | Value |
| --- | --- |
| Total chest radiography examinations | 298,991 |
| Total patients | 114,866 |
| Age, years, median (range) | 45 (16-91) |
| Sex |  |
| Male | 61,447 (53.5%) |
| Female | 53,419 (46.5%) |
| Smoking history |  |
| Current smoker | 21,837 (19%) |
| Former smoker | 23,458 (20.4%) |
| Never smoker | 69,405 (60.4%) |
| Unknown | 166 (0.1%) |
| Number of screening examinations per patient |  |
| 1 time | 44,160 (38.4%) |
| 2 times | 20,483 (17.8%) |
| 3 times | 13,305 (11.6%) |
| 4 times | 12,541 (10.9%) |
| 5 times | 22,618 (19.7%) |
| 6 times or more | 1,759 (1.5%) |
| Confirmed lung cancer, n (%) | 48 (0.04%) |
| Examinations by year |  |
| 2019 | 60,363 |
| 2020 | 52,757 |
| 2021 | 60,769 |
| 2022 | 62,092 |
| 2023 | 63,010 |

**Supplementary Table 2.** Detailed annual 2 x 2 data for the all-score analysis. Year-by-year contingency tables for the primary all-score analysis, including the numbers of true-positive, false-positive, false-negative, and true-negative examinations, together with annual sensitivity, specificity, positive predictive value, and negative predictive value. TP; true-positive, FP; false-positive, FN; false-negative, TN; true-negative, PPV; positive predictive value, NPV; negative predictive value, Cl; confidence intervals..

| Year | TP | FP | FN | TN | Sensitivity (%; 95% CI) | Specificity (%; 95% CI) | PPV (%; 95% CI) | NPV (%; 95% CI) |
| --- | --- | --- | --- | --- | --- | --- | --- | --- |
| 2019 | 1,267 | 12,211 | 452 | 46,433 | 73.7 (71.6–75.8) | 79.2 (78.9–79.5) | 9.4 (8.9–9.9) | 99.0 (98.9–99.1) |
| 2020 | 1,084 | 10,498 | 411 | 40,764 | 72.5 (70.0–74.8) | 79.5 (79.2–79.9) | 9.4 (8.9–9.9) | 99.0 (98.9–99.1) |
| 2021 | 1,153 | 12,091 | 467 | 47,058 | 71.2 (68.9–73.5) | 79.6 (79.2–79.9) | 8.7 (8.2–9.2) | 99.0 (98.9–99.1) |
| 2022 | 1,217 | 12,554 | 481 | 47,840 | 71.7 (69.5–73.8) | 79.2 (78.9–79.6) | 8.8 (8.3–9.3) | 99.0 (98.9–99.1) |
| 2023 | 1,254 | 11,974 | 507 | 49,275 | 71.2 (69.0–73.2) | 80.5 (80.1–80.8) | 9.5 (9.0–10.0) | 99.0 (98.9–99.1) |
| Overall | 5,975 | 59,328 | 2,318 | 231,370 | 72.0 (70.4–73.5) | 79.6 (79.3–79.8) | 9.1 (8.8–9.5) | 99.0 (98.9–99.1) |

**Supplementary Table 3.** Annual radiologist-referenced performance metrics from 2019 to 2023 for the all-score analysis Year Examinations Sensitivity. (%, **95% Cl) Specificity** (%, **95% Cl) PPV** (%, **95% Cl) NPV** (%, **95% Cl)** Year-by-year diagnostic performance of the Al system from 2019 through 2023 in the all-score analysis, including the number of included examinations and corresponding sensitivity, specificity, PPV, and NPV. PPV; positive predictive value, NPV; negative predictive value, Cl; confidence intervals.

| Year | Examinations | Sensitivity (%; 95% CI) | Specificity (%; 95% CI) | PPV (%; 95% CI) | NPV (%; 95% CI) |
| --- | --- | --- | --- | --- | --- |
| 2019 | 60,363 | 73.7 (71.6–75.8) | 79.2 (78.9–79.5) | 9.4 (8.9–9.9) | 99.0 (98.9–99.1) |
| 2020 | 52,757 | 72.5 (70.0–74.8) | 79.5 (79.2–79.9) | 9.4 (8.9–9.9) | 99.0 (98.9–99.1) |
| 2021 | 60,769 | 71.2 (68.9–73.5) | 79.6 (79.2–79.9) | 8.7 (8.2–9.2) | 99.0 (98.9–99.1) |
| 2022 | 62,092 | 71.7 (69.5–73.8) | 79.2 (78.9–79.6) | 8.8 (8.3–9.3) | 99.0 (98.9–99.1) |
| 2023 | 63,010 | 71.2 (69.0–73.2) | 80.5 (80.1–80.8) | 9.5 (9.0–10.0) | 99.0 (98.9–99.1) |
| Total | 298,991 | 72.0 (70.4–73.5) | 79.6 (79.3–79.8) | 9.1 (8.8–9.5) | 99.0 (98.9–99.1) |

Year-by-year diagnostic performance of the AI system from 2019 through 2023 in the all-score analysis, including the number of included examinations and corresponding sensitivity, specificity, PPV, and NPV. PPV; positive predictive value, NPV; negative predictive value, CI; confidence intervals.

**Supplementary Table 4.** Discordant Al-positive patterns in the all-score analysis Section A: Routine screening category breakdown. Distribution of routine screening categories and major radiologist-reported findings among Al-positive/routine-normal examinations in the all-score analysis. Discordant Al-positive examinations were defined as Al-positive and routine-normal examinations in the all-score analysis. Section A shows the breakdown of routine screening categories, and section B shows the distribution of major radiologist-reported findings. Percentages are calculated among all discordant Al-positive examinations in the all-score analysis.

| Routine screening category | n | Proportion (%) |
| --- | --- | --- |
| A (no abnormality) | 45111 | 76.0 |
| B (minor abnormality) | 14217 | 24.0 |
| Total | 59328 | 100.0 |

**Section B: Radiologist-reported findings breakdown**
| Radiologist-reported finding | n | Proportion (%) |
| --- | --- | --- |
| No abnormality | 45111 | 76.0 |
| Old inflammatory changes | 12299 | 20.7 |
| Atelectasis | 1207 | 2.0 |
| Pleural thickening | 394 | 0.7 |
| Congenital change | 109 | 0.2 |
| Post-surgery | 48 | 0.1 |
| Pleural calcification | 38 | 0.1 |
| Benign tumor | 20 | 0.0 |
| Cardiovascular System Abnormalities | 12 | 0.0 |
| Chest wall calcification | 12 | 0.0 |
| Cyst | 9 | 0.0 |
| Calcification | 5 | 0.0 |
| Chest wall mass | 3 | 0.0 |
| Old tuberculosis | 2 | 0.0 |
| Interstitial pneumonia | 1 | 0.0 |
| Others | 485 | 0.8 |
| Total | 59328 | 100.0 |

Distribution of routine screening categories and major radiologist-reported findings among AI-positive/routine-normal examinations in the all-score analysis. Discordant AI-positive examinations were defined as AI-positive and routine-normal examinations in the all-score analysis. Section A shows the breakdown of routine screening categories, and section B shows the distribution of major radiologist-reported findings. Percentages are calculated among all discordant AI-positive examinations in the all-score analysis.

**Supplementary Table 5.** Detailed annual 2 x 2 data for the nodule-focused analysis. Year-by-year contingency tables for the secondary nodule-focused analysis, including the numbers of true-positive, false-positive, false-negative, and true-negative examinations, together with annual sensitivity, specificity, positive predictive value, and negative predictive value. An overall summary across the full study period is also shown. TP; true-positive, FP; false-positive, **FN;** false-negative, **TN;** true-negative, PPV; positive predictive value, and NPV; negative predictive value, CI; confidence intervals..

| Year | TP | FP | FN | TN | Sensitivity (%; 95% CI) | Specificity (%; 95% CI) | PPV (%; 95% CI) | NPV (%; 95% CI) |
| --- | --- | --- | --- | --- | --- | --- | --- | --- |
| 2019 | 91 | 5074 | 14 | 55184 | 86.7 (80.5–93.0) | 91.6 (91.4–91.8) | 1.8 (1.4–2.1) | 100.0 (100.0–100.0) |
| 2020 | 73 | 4279 | 7 | 48398 | 91.2 (84.8–97.1) | 91.9 (91.7–92.1) | 1.7 (1.3–2.1) | 100.0 (100.0–100.0) |
| 2021 | 66 | 4783 | 8 | 55912 | 89.2 (81.2–96.0) | 92.1 (91.9–92.3) | 1.4 (1.0–1.7) | 100.0 (100.0–100.0) |
| 2022 | 74 | 4844 | 15 | 57159 | 83.1 (75.0–90.7) | 92.2 (92.0–92.4) | 1.5 (1.2–1.9) | 100.0 (100.0–100.0) |
| 2023 | 94 | 5422 | 15 | 57479 | 86.2 (79.8–92.7) | 91.4 (91.2–91.6) | 1.7 (1.4–2.1) | 100.0 (100.0–100.0) |
| Overall | 398 | 24402 | 59 | 274132 | 87.1 (84.1–90.5) | 91.8 (91.7–92.0) | 1.6 (1.4–1.8) | 100.0 (100.0–100.0) |

**Supplementary Table 6.** Discordant Al-positive patterns in the nodule-focused analysis. Distribution of routine screening categories and major radiologist-reported findings among examinations classified as discordant Al-positive in the nodule-focused analysis. Discordant Al-positive examinations were defined as Al-positive and radiologist-negative examinations in the nodule-focused analysis. Section A shows the breakdown of routine screening categories, and section B shows the distribution of major radiologist-reported findings together with the mean nodule score for each finding category. Percentages are calculated among all discordant Al--positive examinations in the nodule-focused analysis.

| Routine screening category | n | Proportion (%) |
| --- | --- | --- |
| A (no abnormality) | 15673 | 64.2 |
| B (minor abnormality) | 6166 | 25.3 |
| C (repeat examination or interval follow-up) | 1993 | 8.2 |
| D (detailed diagnostic evaluation or treatment) | 374 | 1.5 |
| E (Under treatment) | 196 | 0.8 |
| Total | 24402 | 100.0 |

**Section B: Radiologist-reported findings breakdown**
| Radiologist-reported finding | n | Proportion (%) | Mean nodule score |
| --- | --- | --- | --- |
| No abnormality | 15673 | 61.7 | 26.9 |
| Old inflammatory changes | 6772 | 26.6 | 32.1 |
| Cyst | 659 | 2.6 | 27.4 |
| Post-surgery | 572 | 2.3 | 28.8 |
| Non-tuberculosis mycobacterium | 186 | 0.7 | 46.4 |
| Atelectasis | 180 | 0.7 | 31.5 |
| Pleural thickening | 158 | 0.6 | 31.7 |
| Interstitial pneumonia | 137 | 0.5 | 28.4 |
| Bronchiectasis | 118 | 0.5 | 36.3 |
| Pneumonia | 84 | 0.3 | 47.0 |
| Cardiovascular System Abnormalities | 79 | 0.3 | 40.0 |
| Mediastinal mass | 68 | 0.3 | 38.1 |
| Emphysematous change | 61 | 0.2 | 30.3 |
| Congenital change | 58 | 0.2 | 37.2 |
| Sarcoidosis | 38 | 0.1 | 35.8 |
| Pleural calcification | 33 | 0.1 | 47.0 |
| Chest wall mass | 30 | 0.1 | 64.2 |
| Benign tumor | 18 | 0.1 | 43.8 |
| Old tuberculosis | 16 | 0.1 | 66.2 |
| Pleural effusion | 15 | 0.1 | 36.4 |
| Pneumothorax | 12 | 0.0 | 31.2 |
| Thyroid mass | 12 | 0.0 | 42.6 |
| Chest wall calcification | 10 | 0.0 | 70.8 |
| Calcification | 6 | 0.0 | 62.9 |
| Thyroid goiter | 4 | 0.0 | 22.7 |
| Tuberculosis | 2 | 0.0 | 40.6 |
| Others | 419 | 1.6 | 49.0 |
| Total | 25420 | 100.0 | 29.3 |

**Supplementary Table 7.** ROG-derived operating points and comparison with the prespecified operational threshold. Comparison of the prespecified manufacturer-recommended operating threshold and the receiver operating characteristic (ROC)-derived Youden-optimal threshold for the all-score and nodule-focused analyses. In the all-score analysis, Al positivity was defined as a score at or above the specified threshold in any of the ten Al output categories, and the reference framework was routine radiologist judgement of abnormality defined as categories other than A or B. In the nodule-focused analysis, AI positivity was defined as a nodule or mass score at or above the specified threshold, and the reference framework was radiologist-reported pulmonary nodule or mass findings. Sensitivity and specificity are reported relative to the predefined routine radiologist reference framework. The Youden index was calculated as sensitivity+ specificity - 1. AUC, area under the receiver operating characteristic curve; Cl, confidence interval.

| Analysis | Operating point | Threshold | Sensitivity (%) | Specificity (%) | Youden index | AUC (95% CI) |
| --- | --- | --- | --- | --- | --- | --- |
| All-score | Prespecified threshold | 15.0 | 72.0 | 79.6 | 0.516 | 0.828 (0.823–0.833) |
|  | Youden-optimal threshold | 16.9 | 71.2 | 80.8 | 0.520 | — |
| Nodule-focused | Prespecified threshold | 15.0 | 87.1 | 91.8 | 0.789 | 0.950 (0.938–0.962) |
|  | Youden-optimal threshold | 18.1 | 85.3 | 94.0 | 0.793 | — |

**Supplementary Table 8.** Summary of confirmed lung cancer cases and retrospective positivity patterns. Summary of confirmed lung cancer cases identified in the linked outcome cohort and their detection patterns according to the relationship between Al positivity and radiologist positivity at relevant screening timepoints. Summary statistics for lead time to confirmed diagnosis are also shown for cases with evaluable timelines.

| **Retrospective positivity pattern** | **N** | **%** |
| --- | --- | --- |
| Earlier AI positivity | 11 | 22.9 |
| AI-only positivity before diagnosis | 10 | 20.8 |
| Radiologist-only positivity | 2 | 4.2 |
| Simultaneous AI and radiologist positivity | 20 | 41.7 |
| No AI or radiologist positivity at relevant timepoint | 5 | 10.4 |

**Supplementary Table 9.** Case-level timeline data for confirmed lung cancer cases. Case-level characteristics and screening timelines for 48 confirmed lung cancer cases in the linked outcome cohort. The table includes demographic characteristics, screening history, timing of the first Al-positive and first radiologist-positive examinations, detection category, and tumour characteristics. Day O was defined as the date of diagnostic radiographic abnormality, according to available clinical records. Earlier events are shown as negative values. Retrospective positivity categories were defined according to the timing of Al positivity and routine radiologist positivity. Retrospective positivity categories were defined as simultaneous Al and radiologist positivity, earlier Al positivity, Al-only positivity, radiologist-only positivity, or no Al or radiologist positivity at the relevant timepoint. Tumour stage was assigned according to the 9th edition of the TNM Classification for Lung Cancer. NA indicates not available.

| Case ID | Age, years | Sex | Total screening examinations | Pre-diagnostic screening examinations | First screening to day 0, days | First AI-positive result to day 0, days | First radiologist-positive result to day 0, days | Retrospective positivity category | Interval from first AI positivity to diagnosis (days) | Pathology | Stage (TNM, 9th edition) |
| --- | --- | --- | --- | --- | --- | --- | --- | --- | --- | --- | --- |
| 1 | 66 | M | 3 | 0 | 0 | 0 | 0 | Simultaneous AI and radiologist positivity | 0 | Lung Cancer | NA |
| 2 | 57 | M | 4 | 1 | -373 | 0 | 0 | Simultaneous AI and radiologist positivity | 0 | Squamous cell lung cancer | NA |
| 3 | 58 | M | 6 | 4 | -1409 | -1409 | 0 | Earlier AI positivity | 1409 | Lung Cancer | NA |
| 4 | 70 | M | 3 | 0 | 0 | 0 | NA | AI-only positivity before diagnosis | 0 | Lung Cancer | NA |
| 5 | 68 | M | 4 | 3 | -1461 | 0 | 0 | Simultaneous AI and radiologist positivity | 0 | Lung Cancer | NA |
| 6 | 63 | F | 2 | 1 | -717 | -717 | 0 | Earlier AI positivity | 717 | Lung Adenocarcinoma | IB |
| 7 | 58 | F | 5 | 0 | 0 | 0 | 0 | Simultaneous AI and radiologist positivity | 0 | Lung Adenocarcinoma | IIIA |
| 8 | 53 | M | 5 | 1 | -365 | -365 | 0 | Earlier AI positivity | 365 | Lung Adenocarcinoma | IIB |
| 9 | 47 | F | 4 | 1 | -364 | -364 | 0 | Earlier AI positivity | 364 | Lung Adenocarcinoma | IB |
| 10 | 69 | M | 5 | 1 | -370 | -370 | NA | AI-only positivity before diagnosis | 370 | Lung Cancer | NA |
| 11 | 55 | F | 4 | 2 | -679 | -679 | 0 | Earlier AI positivity | 679 | Lung Adenocarcinoma | IVB |
| 12 | 58 | M | 4 | 3 | -1431 | NA | NA | No AI or radiologist positivity at relevant timepoint | NA | Lung Cancer | NA |
| 13 | 61 | M | 4 | 1 | -354 | 0 | 0 | Simultaneous AI and radiologist positivity | 0 | Lung Adenocarcinoma | IA2 |
| 14 | 59 | F | 2 | 1 | -357 | -357 | 0 | Earlier AI positivity | 357 | Lung Cancer | NA |
| 15 | 50 | M | 4 | 2 | -798 | -798 | NA | AI-only positivity before diagnosis | 798 | Lung Adenocarcinoma | IIB |
| 16 | 67 | F | 4 | 1 | -379 | -379 | 0 | Earlier AI positivity | 379 | Lung Adenocarcinoma | NA |
| 17 | 65 | M | 4 | 3 | -1445 | 0 | NA | AI-only positivity before diagnosis | 0 | Lung Cancer | NA |
| 18 | 68 | M | 4 | 3 | -1408 | -1408 | 0 | Earlier AI positivity | 1408 | Lung Cancer | NA |
| 19 | 54 | M | 5 | 3 | -959 | -234 | NA | AI-only positivity before diagnosis | 234 | Lung Cancer | NA |
| 20 | 43 | M | 1 | 0 | 0 | 0 | 0 | Simultaneous AI and radiologist positivity | 0 | Lung Adenocarcinoma | IB |
| 21 | 74 | M | 5 | 4 | -1775 | 0 | 0 | Simultaneous AI and radiologist positivity | 0 | Lung Adenocarcinoma | IA3 |
| 22 | 61 | M | 2 | 2 | -670 | NA | NA | No AI or radiologist positivity at relevant timepoint | NA | Lung Adenocarcinoma | IV |
| 23 | 63 | M | 5 | 3 | -1093 | -677 | 0 | Earlier AI positivity | 677 | Lung Adenocarcinoma | NA |
| 24 | 53 | M | 3 | 2 | -728 | NA | 0 | Radiologist-only positivity | NA | Lung Adenocarcinoma | IIIA |
| 25 | 66 | M | 4 | 2 | -671 | -377 | NA | AI-only positivity before diagnosis | 377 | Lung Adenocarcinoma | IB |
| 26 | 44 | F | 4 | 0 | 0 | 0 | 0 | Simultaneous AI and radiologist positivity | 0 | Lung Adenocarcinoma | IA2 |
| 27 | 61 | M | 4 | 1 | -391 | -391 | -391 | Simultaneous AI and radiologist positivity | 0 | Lung Cancer | NA |
| 28 | 58 | F | 3 | 2 | -1122 | 0 | 0 | Simultaneous AI and radiologist positivity | 0 | Lung Adenocarcinoma | IA2 |
| 29 | 59 | M | 4 | 1 | -370 | 0 | 0 | Simultaneous AI and radiologist positivity | 0 | Lung Cancer | NA |
| 30 | 49 | M | 4 | 3 | -1336 | -701 | NA | AI-only positivity before diagnosis | 701 | Lung Adenocarcinoma | NA |
| 31 | 57 | M | 4 | 1 | -351 | -351 | 0 | Earlier AI positivity | 351 | Lung Adenocarcinoma | IB |
| 32 | 61 | M | 1 | 0 | 0 | NA | NA | No AI or radiologist positivity at relevant timepoint | NA | Lung Cancer | NA |
| 33 | 45 | M | 1 | 0 | 0 | NA | NA | No AI or radiologist positivity at relevant timepoint | NA | Lung Adenocarcinoma | IA1 |
| 34 | 62 | M | 1 | 0 | 0 | 0 | 0 | Simultaneous AI and radiologist positivity | 0 | Squamous cell lung cancer | IIIC |
| 35 | 52 | F | 1 | 0 | 0 | 0 | 0 | Simultaneous AI and radiologist positivity | 0 | Lung Adenocarcinoma | IA3 |
| 36 | 69 | M | 3 | 2 | -779 | -779 | NA | AI-only positivity before diagnosis | 779 | Lung Cancer | NA |
| 37 | 44 | M | 4 | 2 | -1092 | 0 | 0 | Simultaneous AI and radiologist positivity | 0 | Lung Adenocarcinoma | NA |
| 38 | 51 | F | 3 | 2 | -752 | -315 | NA | AI-only positivity before diagnosis | 315 | Lung Adenocarcinoma | IVB |
| 39 | 64 | M | 4 | 0 | 0 | 419 | NA | No AI or radiologist positivity at relevant timepoint | NA | Lung Cancer | NA |
| 40 | 67 | F | 2 | 0 | 0 | 0 | 0 | Simultaneous AI and radiologist positivity | 0 | Lung Cancer | NA |
| 41 | 70 | M | 1 | 1 | -315 | -315 | NA | AI-only positivity before diagnosis | 315 | Lung Adenocarcinoma | IVA |
| 42 | 64 | M | 2 | 2 | -709 | -709 | -368 | Earlier AI positivity | 341 | Lung Cancer | NA |
| 43 | 68 | M | 1 | 0 | 0 | 0 | 0 | Simultaneous AI and radiologist positivity | 0 | Lung Adenocarcinoma | IVA |
| 44 | 70 | M | 1 | 0 | 0 | 0 | 0 | Simultaneous AI and radiologist positivity | 0 | Lung Cancer | IVA |
| 45 | 71 | M | 1 | 0 | 0 | 0 | 0 | Simultaneous AI and radiologist positivity | 0 | Lung Adenocarcinoma | IVB |
| 46 | 63 | F | 1 | 0 | 0 | 0 | 0 | Simultaneous AI and radiologist positivity | 0 | Lung Adenocarcinoma | IA1 |
| 47 | 64 | F | 4 | 1 | -445 | NA | 0 | Radiologist-only positivity | NA | Lung Adenocarcinoma | IA2 |
| 48 | 58 | M | 2 | 1 | -318 | 0 | 0 | Simultaneous AI and radiologist positivity | 0 | Lung Adenocarcinoma | IVA |

**Supplementary Figure 1.**
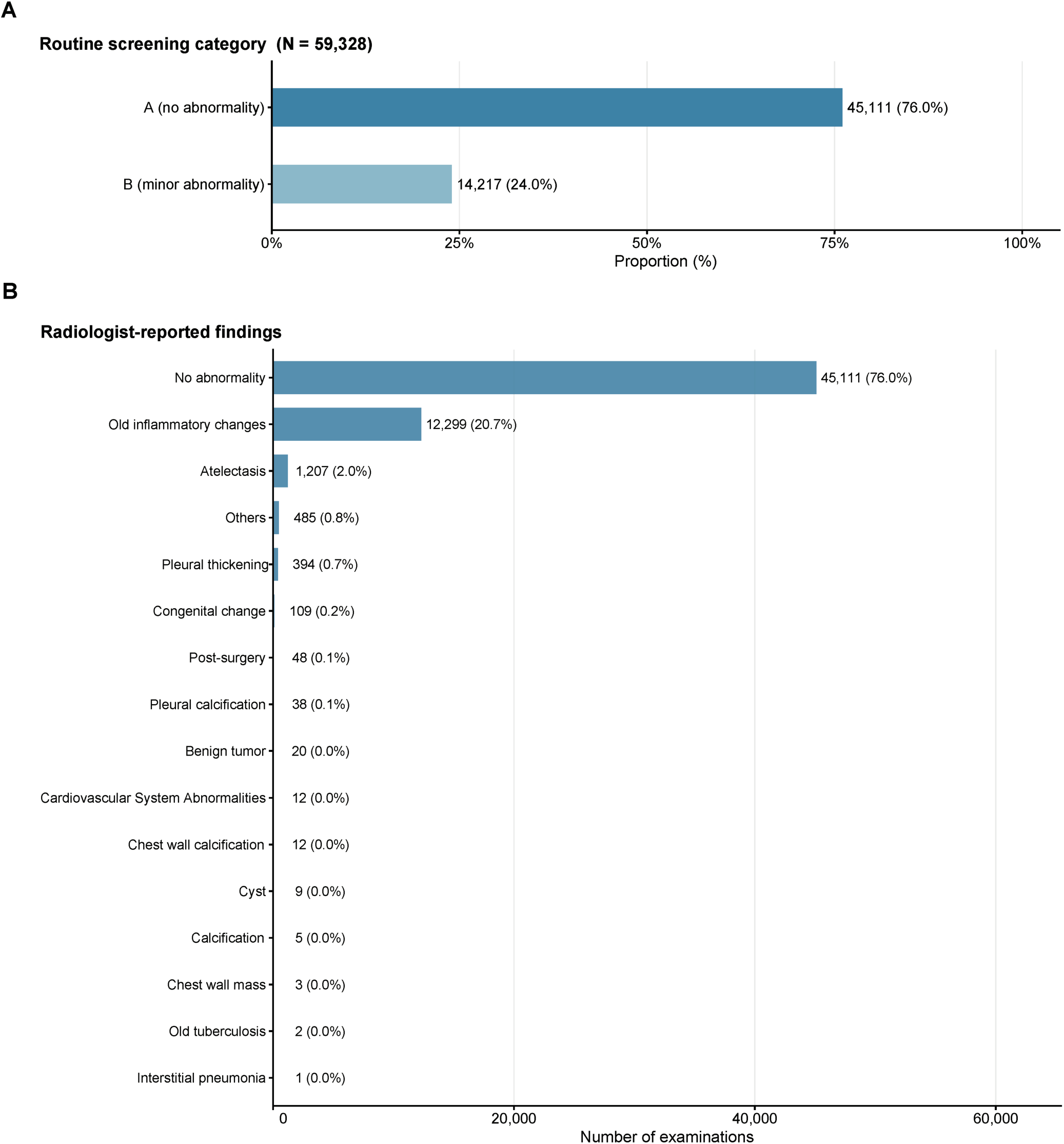
Breakdown of discordant Al-positive examinations in the primary all-score analysis. Routine screening categories and major radiologist-reported findings among discordant Al-positive examinations in the primary all-score analysis. The upper panel shows the distribution of routine screening categories **(N** = 59,328), and the lower panel shows the top 10 radiologist-reported findings by frequency. Counts and, where appropriate, proportions (%) are shown. Al positivity in the primary all-score analysis was defined as a score of 15 or higher in any of the 10 thoracic abnormality categories.

**Supplementary Figure 2.**
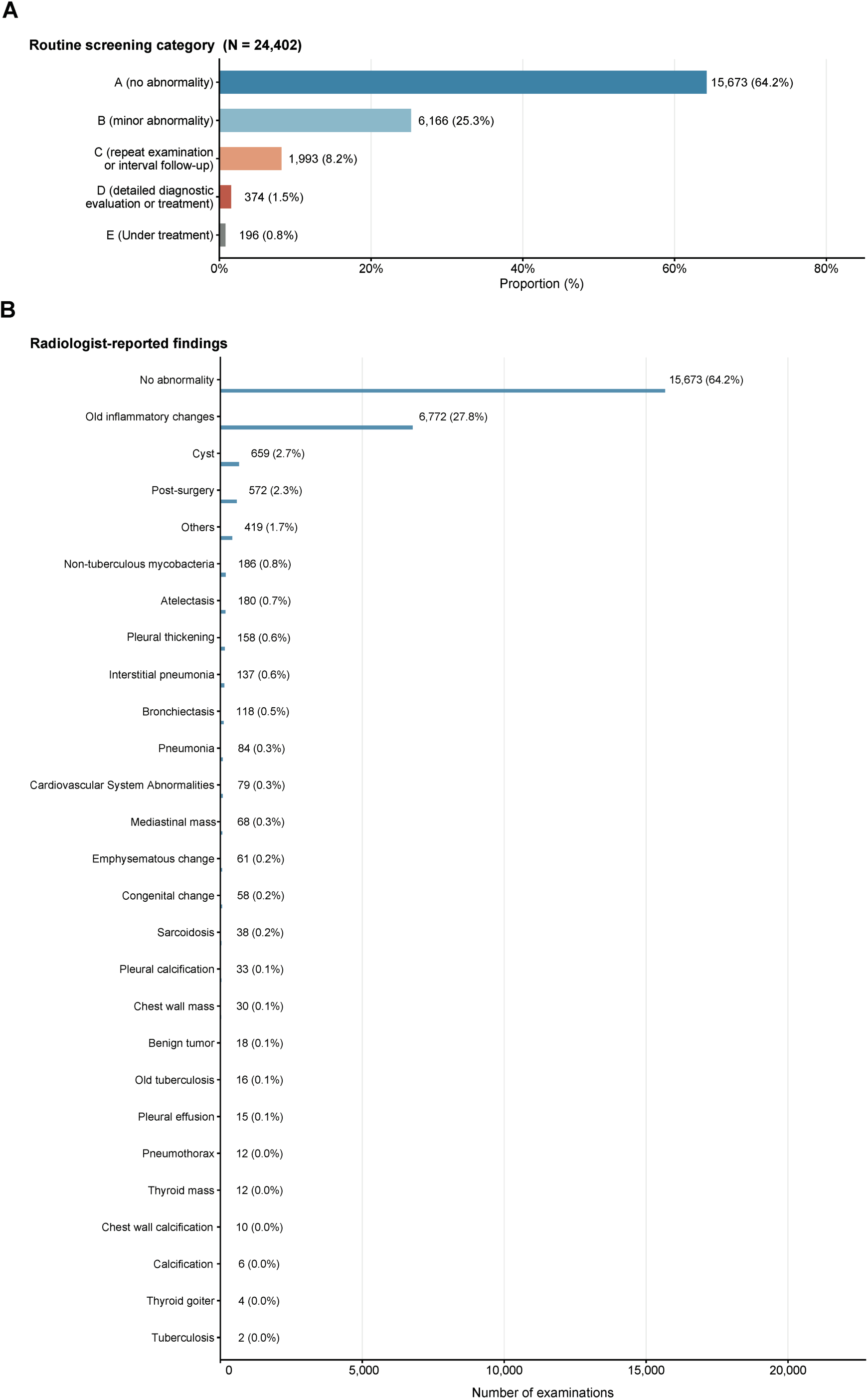
Breakdown of discordant Al-positive examinations in the secondary nodule-focused analysis. Routine screening categories and major radiologist-reported findings among discordant Al-positive examinations in the secondary nodule-focused analysis. The upper panel shows the distribution of routine screening categories **(N** = 24,402), and the lower panel shows the top 10 radiologist-reported findings by frequency. Counts and, where appropriate, proportions (%) are shown. Al positivity in the secondary nodule-focused analysis was defined as a nodule or mass score of 15 or higher.

**Supplementary Figure 3.**
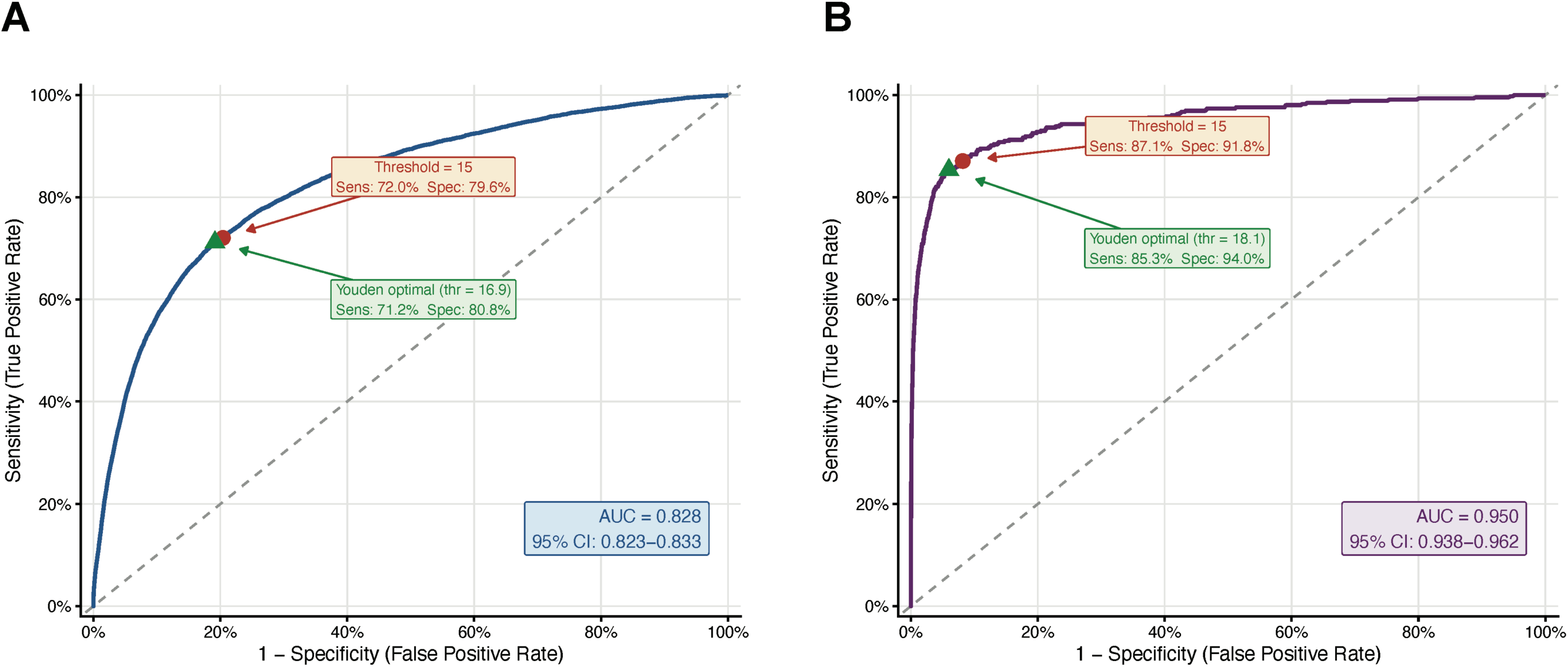
Receiver operating characteristic analyses and operating threshold evaluation for the all-score and nodule-focused analyses. Receiver operating characteristic (ROC) curves demonstrating the performance of the Al system across the full range of operating thresholds for the all-score analysis **(A)** and the nodule-focused analysis (**8**). In the all-score analysis, the reference framework was routine radiologist-defined abnormality, defined as health check-up categories other than A or B. In the nodule-focused analysis, the reference framework was radiologist-reported pulmonary nodule or mass findings. The red circle indicates the prespecified manufacturer-recommended operational threshold (score ≥ 15) used throughout the study, whereas the green triangle indicates the Youden-optimal threshold identified by maximizing the Youden index (sensitivity+ specificity - 1). The dashed diagonal line represents the line of no discrimination. Area under the ROC curve (AUC) values and corresponding 95% confidence intervals (Cls) are shown for each analysis. The proximity of the prespecified threshold to the ROG-derived optimal threshold supports the use of the selected operating point for implementation-oriented evaluation.

### Supplementary Materials and Methods

#### Ethics, consent, and data governance

This study was approved by the Institutional Review Board of The Jikei University School of Medicine (approval number 35-386 [12023]). The requirement for written informed consent was waived because of the retrospective use of de-identified data, and an opt-out approach was used in accordance with institutional policy. Data linkage and analysis were performed using anonymised identifiers in a secure research environment, and no directly identifiable information was available to the study investigators.

#### Participants

Eligible examinations were screening chest radiographs acquired during the study period. The source dataset contained 299,745 examinations from 114,974 individuals. Records were linked to AI outputs using anonymised identifier and examination date. A total of 299,712 examinations were successfully linked. Examinations with missing values in one or more AI score fields were excluded, yielding a final cohort of 298,991 examinations from 114,866 patients. The selection flow and exclusion criteria were prespecified and applied consistently across years.

#### AI system and index test

The index test was a commercially available deep learning-based AI system for chest radiography (CXR-AID version 3.0; FUJIFILM, Tokyo, Japan). The system was developed using a large chest radiography dataset annotated by board-certified radiologists and underwent software verification and validation as part of the manufacturer’s medical device development process. In addition, the software has received regulatory approval in Japan based on dedicated reader-performance studies conducted for regulatory evaluation. The AI system outputs localisation maps and continuous abnormality scores ranging from 0 to 100, representing the estimated probability of abnormality for ten thoracic findings: nodule or mass, infiltration, pneumothorax, atelectasis, calcification, scar, pleural effusion, pneumoperitoneum (free air), cardiomegaly, and mediastinal widening. The AI analysis was performed as an automated post-processing step on stored chest radiographs and did not modify the original routine radiologist judgement used as the reference framework in this retrospective evaluation. To support transparent reporting of the index test, we prespecified both the operating threshold and the output categories used in each analysis (1, 2). For the primary analysis, AI positivity was defined as a score of 15 or higher in any of the ten output categories (all-score analysis). For the secondary analysis, AI positivity was defined as a nodule or mass score of 15 or higher (nodule-focused analysis). The threshold of 15 was applied consistently across the study dataset because this manufacturer-recommended threshold was prespecified for evaluation of the commercial software.

#### Definitions for the nodule-focused analysis

Radiologist-positive nodule examinations were defined by the presence of keywords corresponding to pulmonary nodule or mass findings in the routine radiology text fields. Only pulmonary field abnormalities were counted as positive. Findings involving the chest wall, mediastinum, thyroid, neck, benign tumours, or hilar enlargement were treated as negative in this lesion-specific analysis. This definition was selected to align the reference classification as closely as possible with the target condition of pulmonary nodules or masses in the screening setting.

#### Statistical analysis

We constructed 2 × 2 tables for the all-score and nodule-focused analyses. Sensitivity, specificity, PPV, and NPV were calculated using standard definitions, but these metrics were interpreted as radiologist-referenced operational concordance metrics because routine radiologist judgement, rather than universal CT or pathologic verification, served as the reference framework. For clarity, true-positive, false-positive, false-negative, and true-negative classifications refer to concordance or discordance with the predefined routine reference framework and do not necessarily represent biological truth or lung cancer status. Discordant review volume was defined as the number of examinations in which AI output and routine radiologist judgement were not concordant, including AI-positive/radiologist-negative and AI-negative/radiologist-positive examinations. Analyses were descriptive and focused on implementation-relevant features of AI deployment, including positivity rates, discordant volumes, score-band distributions, and outcome-linked patterns in lung cancer cases. In line with reporting guidance for observational and diagnostic accuracy studies, we explicitly report cohort selection, index test definition, reference framework, and primary diagnostic metrics (1–3).

#### Confidence interval estimation accounting for repeated measurements

Because the cohort comprised 298,991 examinations from 114,866 individuals, repeated measurements within the same patient introduce within-person correlation that standard interval methods do not account for. Ninety-five percent confidence intervals for sensitivity, specificity, PPV, and NPV were therefore estimated using a non-parametric cluster bootstrap procedure (B = 1,000 iterations) in which the resampling unit was the individual patient; all examinations from a sampled patient were included in each replicate, thereby preserving the within-person correlation structure. The 2.5th and 97.5th percentiles of the bootstrap distribution were used as interval bounds. The same procedure was applied to the overall cohort and to each calendar year separately. A random seed was fixed to ensure reproducibility. All analyses were conducted in R (version 4.5.1; R Foundation for Statistical Computing, Vienna, Austria).

#### Threshold sensitivity analysis

To assess the robustness of primary findings to the choice of operating threshold, diagnostic performance was evaluated across a range of AI score thresholds in addition to the prespecified threshold of 15. Thresholds of 10, 12, 15, 18, 25, and 50 were evaluated for both the all-score and nodule-focused analyses. For each threshold, sensitivity, specificity, PPV, NPV, and discordant examination volume were computed, with 95% confidence intervals derived from the cluster bootstrap procedure described above. This range was selected to cover clinically plausible operating points from sensitivity-prioritised threshold 10 to specificity-prioritised threshold 50 settings, and includes the manufacturer-recommended threshold 15, the Youden-index optimal threshold identified in receiver operating characteristic analysis (approximately 18), and two higher reference points (25 and 50).

